# Effect of COVID-19 vaccine on long-COVID: A 2-year follow-up observational study from hospitals in north India

**DOI:** 10.1101/2022.07.18.22277740

**Authors:** Sandeep Budhiraja, Abhaya Indrayan, Monica Mahajan

**Author notes:** **Corresponding author** Dr. Sandeep Budhiraja MD, DNB, FACP, FRCP (Edin), Group Medical Director, Max Healthcare, Senior Director, Institute of Internal Medicine, Max Super Speciality Hospital (Unit of Devki Devi Foundation), 2, Press Enclave Road, Saket, New-Delhi-110017, India.

## Abstract

**Introduction:** Long-COVID syndrome encompasses a constellation of fluctuating, overlapping systemic symptoms after COVID. We know that vaccination reduces the risk of hospitalization and death but not of re-infections. How these vaccines impact long-COVID is under debate. The current study was designed to analyze the patterns of long-COVID amongst vaccinated and unvaccinated hospitalized patients during the three waves in India.

**Methods:** The computerized medical records of the patients admitted to a group of hospitals in the National Capital Region of Delhi with a nasopharyngeal swab positive RT-PCR for SARS-CoV-2, during the three distinct COVID-19 waves, were accessed. Because of large numbers, every 3^rd^ case from the data sheet for the wave-1 and wave-2 but all cases admitted during wave-3 were included because of small numbers (total 6676). The selected patients were telephonically contacted in April 2022 for symptoms and their duration of long-COVID and their vaccination status. Of these, 6056 (90.7%) responded. These were divided into fully vaccinated who received both doses of COVID vaccine at least 14 days before admission (913) and unvaccinated at the time of admission (4616). Others and deaths were excluded. “Symptom-weeks” was calculated as the sum of weeks of symptoms in case of two or more symptoms. The statistical significance was tested, and odds ratio (unadjusted and adjusted) were calculated by logistic regression.

**Results:** Nearly 90% of COVID-19 patients reported at least one symptom irrespective of their vaccination status. Almost three-fourths of these had symptoms lasting up to a month but nearly 15% reported a duration a least 4 weeks including 11% even exceeding one year. During wave-3, significantly more vaccinated patients reported short term post-acute sequelae of COVID-19 than did the unvaccinated group. The cases with diabetes and hypertension had higher odds of reporting at least one symptom when the effect of vaccination, age, sex, severity, and length of stay was adjusted. The fully vaccinated cases had reduced length of stay in the hospital and had a milder disease. Most common symptoms reported by both the groups were fatigue (17.0%), insomnia (15.1%) and myalgia (15%). There were significant differences in the duration and the type of long-COVID symptoms across the three waves, and the presence of comorbidities between the vaccinated and the unvaccinated groups but overall no difference could be detected. No significant difference was seen between the cases receiving covishield and covaxin.

**Conclusions:** Nearly 15% reported symptoms of duration exceeding 4 weeks including 11% exceeding one year. There were significant differences in the specific symptoms with some more common in the vaccinated and some others more common in the unvaccinated but overall the vaccination or the type of vaccine did not significantly alter either the incidence or the duration of long COVID.

## Introduction

Since the declaration of coronavirus disease COVID-19 as a global pandemic by the World Health Organisation on 11th March 2020, India have witnessed continued surges and declines in the prevalence of coronavirus cases causing waves. Three distinct waves of the COVID-19 pandemic occurred: wave-1 from March 2020 till December 2020 caused by the wild Wuhan strain, wave-2 from January 2021 till June 2021 caused mainly by Delta strain, and wave-3 from December 2021 till February 2022 caused mainly by Omicron strain. The virus variants have a common lineage but had varying degrees of transmissibility and disease severity than the progenitor virus due to multiple mutations. The mutations also posed a challenge in preventing and treating the infection.

The SARS-CoV2 virus has not only resulted in acute infections in many cases lasting r up to 4 weeks but also ongoing post-COVID-19 signs and symptoms lasting 4 to 12 weeks. A distinct new entity emerged is the post-COVID syndrome wherein the patients had unexplained symptoms even beyond 12 weeks. These maybe ongoing or new set of symptoms. Long-COVID syndrome, as per the National Institute for Health and Care Excellence (NICE) UK guidelines [1] encompasses a constellation of fluctuating, overlapping systemic symptoms both the ongoing symptomatic COVID (4-12 weeks) and post-COVID syndrome (>12 weeks). All these together may be called post-acute sequelae of COVID-19 (PASC) [2]. The terms long-COVID and PASC are inter-changeably used. The magnitude of the problem is huge with far-reaching mental, physical and financial implications. The Office of National Statistics (ONS) has recorded 600,000 patients with long-COVID during the Omicron outbreak in the UK with 5% of healthcare workers reporting symptoms of long-COVID [3].

Vaccines against SARS-CoV2 were developed at a war-footing, got emergency use approval and wide acceptance in a world fighting a losing battle against an unknown enemy. A question to debate has been whether use of these vaccines has any impact on long-COVID. Various studies have shown conflicting results. While most studies show that vaccination prior to getting COVID-19 infection reduces the incidence and severity of long-COVID [,4 -8], some studies have shown no effect [9] to perhaps increasing the risk of getting long -COVID [10].

For the pathophysiological mechanisms involved in the causation of long-COVID, Castanares-Zapatero et al. [11] searched 11 bibliographic databases. Various neurological symptoms were attributed to hypometabolic activity in various cerebral zones, reduced GABA inhibition, neuro-inflammation and brain microstructural modifications and direct injury to olfactory neuronal pathways. The predominant cardiovascular symptoms were explained by pathophysiological mechanisms like persistent macrovascular and microvascular inflammation caused by increased levels of cytokines, circulating endothelial cells, coagulation activation and the role of auto-immunity. Respiratory system involvement was possibly due to persistent inflammation and dysregulated host response of lung repair, increased plasma biomarkers of lung inflammation and fibrosis, persisting inflammation in lungs, mediastinal lymph nodes, spleen and liver and involvement of iron homeostasis disturbances in end-organ damage. The immune system dysregulation could be the result of persistent immune inflammatory response impairing organ functioning, long-lasting phenotypic and functional disorders of lymphocytes, decreased amounts of dendritic cells and persisting alterations of activation markers and result of auto-immunity and tissue components and persistence of SARS-CoV-2 nucleic acids in tissues.

The current study analyses data collected through computerized medical records and telephonic conversations to study the patterns of long-COVID during the three waves amongst vaccinated and unvaccinated patients. These waves also correspond to Wuhan, Delta, and Omicron variants of Corona virus.

## Methods

The study included patients admitted to a group of tertiary care hospitals in the National Capital Region of Delhi. Their clinical records were accessed from electronic medical records of the hospitals. The diagnosis of COVID-19 was made by a positive RT-PCR test for SARS-CoV-2 from nasopharyngeal swab. The patients were admitted over the three waves starting March 2020 (Wave 1: March-December 2020; Wave 2: January – June 2021; Wave 3: December 2021-February 2022). The records had information on their age, sex, length of stay, admission to ICU, severity, and comorbidities. Only those cases with complete data were considered. There were 11730 admissions in wave-1, 3604 in wave-2, and 1565 in wave-3 in the hospitals. Because of large numbers, we randomly selected every 3^rd^ case from the master sheet for wave-1 and wave-2 by systematic sampling and all cases for wave-3 because of smaller numbers admitted in this wave. This yielded a total of 6676 patients. These patients were contacted telephonically for symptoms and their duration and vaccination status for COVID-19. Of these, 6056 (90.7%) responded after clearly understanding the purpose of the survey. They responded to a structured questionnaire which took on an average 10 minutes per call. For the purpose of present study, patients were asked regarding presence of any symptoms after discharge from the hospital, as evidence of post-acute sequelae of COVID-19 (PASC) or long COVID. The terms post-COVID, long COVID and PASC have been used interchangeably. Depending upon the number of symptoms and the duration for which each symptom persisted, we calculated “symptom-week” as the sum of duration of all symptoms. This metric incorporates both the number and the duration of symptoms as both together have implications for the quality of life.

The patients were categorised (depending upon the duration of symptoms), as:

No PASC: no reported symptoms, after discharge from the hospital

Short-term PASC: symptoms with duration between 1-4 weeks after discharge from the hospital

Intermediate-term PASC: Symptoms persisting between 1-5 months post-discharge from the hospital

Long-term PASC: Symptoms persisting for 6 months or more, after discharge from the hospital

The respondents were 3878 (64.0%) admitted in wave-1, 1163 (19.2%) admitted in wave-2, and 1015 (16.8%) admitted in wave-3. The tele-calling was done during second and third weeks of April 2022 and April 15 was considered as the cut-off date for the analysis.

A total of 133 deaths out of 6056 (2.2%) were reported from the discharge to the tele-calling. It was unethical to ask for PASC in these cases and so they were excluded from the analysis, leaving a sample size of 5923. A group of 394 patients was further excluded from the final study results as these patients either had taken one dose of COVID vaccine within 14 days before admission or had taken only one dose of vaccine prior to getting admitted for COVID-19 and hence, were considered unprotected. The flow chart is in Figure 1.

**Figure 1.**
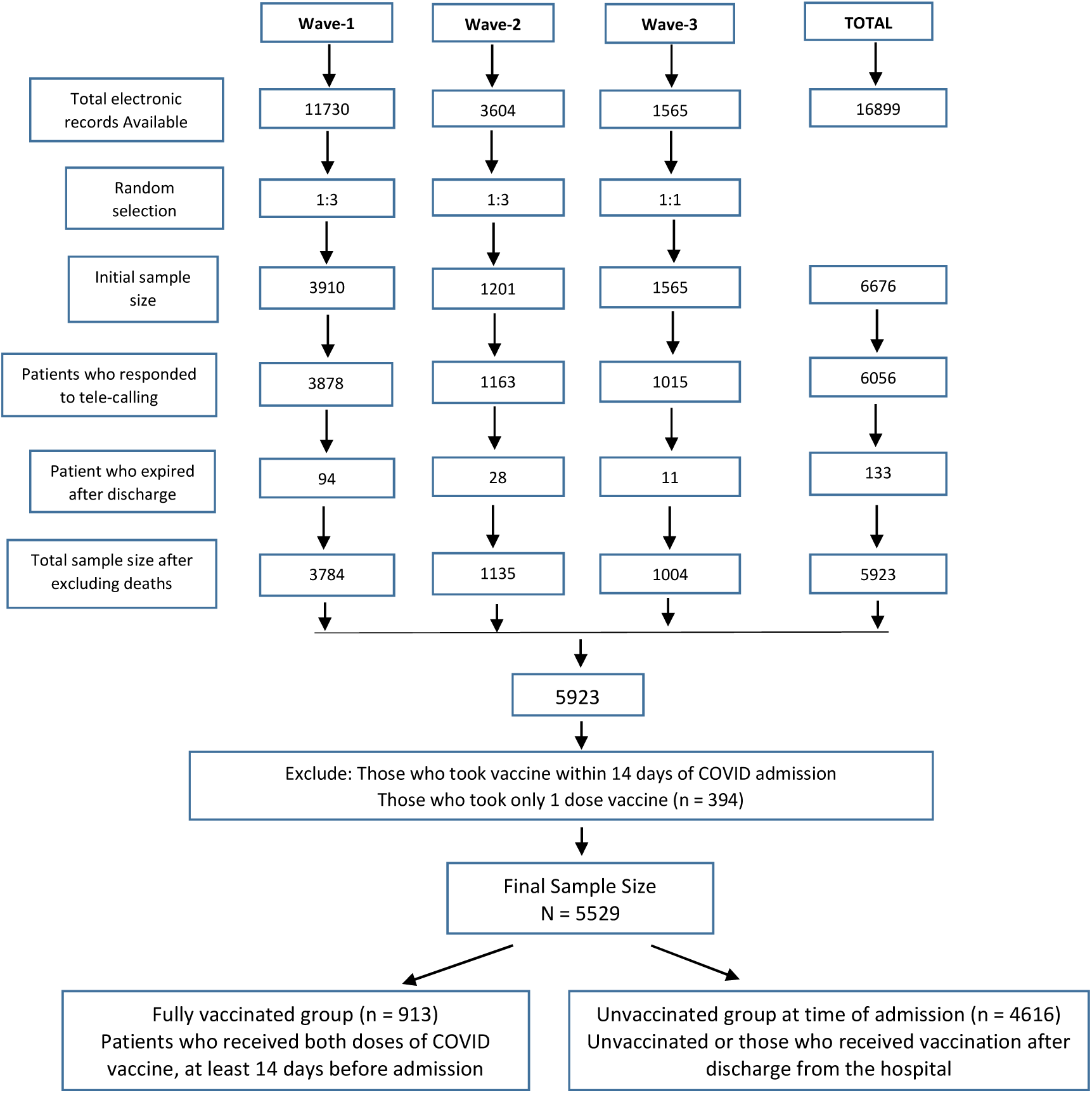
Flow chart of cases.

The final sample size of 5529 patients was divided into 2 groups:

Fully vaccinated group (913) – Patients who received both doses of COVID vaccine at least 14 days before admission

Unvaccinated at the time of admission (4616) – Unvaccinated or those who received vaccination after discharge from the hospital

The two vaccines available and approved for use in India during this period were the recombinant adenovirus vector vaccine by AstraZeneca ChAdOx1nCoV-19 (Available as Covishield in India, manufactured by Serum Institute of India Pvt Ltd) and a whole-virion inactivated vero cell derived vaccine (available as Covaxin in India, manufactured by Bharat Biotech). The patients discharged in 2020 (wave-1) were 100% unvaccinated, as the vaccine drive in India started only in January 2021. The patients discharged during wave-2 included both vaccinated and unvaccinated group, and the patients discharged during wave-3 were largely vaccinated but had some unvaccinated cases also.

The severity of COVID-19 was classified as mild, moderate, and severe, as per the criteria of Ministry of Health and Family Welfare, Govt. of India [12].

Since all these patients were discharged from hospitals over varying periods of time between 2020 till Feb 2022, the periods of follow-up varied from 2 months (for those discharged in February 2022 wave-3) to as much as 2 years for those discharged in March 2020 (wave-1). A larger proportion of the unvaccinated cases were from those who were discharged in 2020 and early 2021 and thus their follow-up period was longer. Vaccinated group was largely contributed by patients discharged in 2022 and some were from those discharged in 2021. The average duration of follow-up for cases admitted in different waves and the minimum and maximum are in Table 1.

**Table 1.** Follow-up period (weeks) in the vaccinated and the unvaccinated cases in the different waves.

| Vaccination status | Wave-1 |  |  | Wave-2 |  |  | Wave-3 |  |  |
| --- | --- | --- | --- | --- | --- | --- | --- | --- | --- |
|  | Mean | Min | Max | Mean | Min | Max | Mean | Min | Max |
| **Vaccinated** | No case |  |  | 52.7 | 51.0 | 55.4 | 12.6 | 5.6 | 22.9 |
| **Unvaccinated** | 83.1 | 67.4 | 106.3 | 56.9 | 50.4 | 71.6 | 12.8 | 6.9 | 16.6 |
| **Total** | 83.1 | 67.4 | 106.3 | 56.8 | 50.4 | 71.6 | 12.6 | 5.6 | 22.9 |

### Statistical analysis

The patients in the vaccinated (Group-1) and the unvaccinated (Group-2) groups were compared by chi-square test for qualitative characteristics such as disease severity and by Student t-test for quantitative characteristics such as age. A multivariable logistic regression was done to find the odds ratio of reporting of symptoms in the vaccinated group vs. the unvaccinated group after adjusting for covariates such as age, sex, length of stay, disease severity, and comorbidities. Symptoms were continuing in some cases at the time of the survey. Symptoms-weeks were calculated as the sum of duration of all symptoms in a person because both the duration and the number of symptoms affect the quality of life. The hazard ratio for symptom-weeks was obtained by Cox regression after adjusting for the covariates. Because of multiple use of the data and in view of a large sample, the significance level was kept at 1% and SPSS 22 was used for calculation as advised [13].

### Ethics Committee Approval and Patient consent

The study titled “Effect of COVID-19 vaccine on long-COVID: a 2-year follow-up observational study from North Indian Hospitals” was approved by the Institutional Ethics Committee, Devki Devi Foundation; address: service floor, office of Ethics Committee, East Block, Next to Conference Room, Max Super Speciality Hospital, Saket (A unit of Devki Devi Foundation), 2, Press Enclave Road, Saket, New Delhi – 110017 vide ref. no. BHR/RS/MSSH/DDF/SKT-2/IEC/IM/22-12, dated 5^th^ July 2022.

## Results

### Baseline characteristics of the cases in the study

On average, the vaccinated patients were older, particularly of age 75 years or more (22.6%) against only 9.1% in the unvaccinated group (Table 2). Females were significantly (P < 0.001) more in the vaccinated group (41.2% vs. 34.3%). Vaccinated patients stayed in the hospital for shorter period (median 5.0 days vs. 7.4 days) – nearly half (45.7%) stayed for less than 5 days against 18.8% of the unvaccinated. ICU admission rate was, however, not much different (nearly 25% in both the groups). There were higher percentage of vaccinated patients with mild COVID-19 at the time of hospitalisation and fewer with moderate severity compared to the unvaccinated patients. The percentage with severe COVID-19 at admission (nearly 28%) was nearly the same in both the groups. Vaccinated group had significantly less patients with diabetes and hypertension but more with dyslipidaemia and hypothyroidism (Table 2).

**Table 2.** Basic characteristics of the patients in the two groups.

| Characteristics | Total |  | Group |  |  |  | P-value |
| --- | --- | --- | --- | --- | --- | --- | --- |
|  |  |  | Vaccinated |  | Unvaccinated |  |  |
|  | No. | Col % | No. | Col % | No. | Col % |  |
| Total no. of patients | 5529 | 100.00% | 913 | 16.50% | 4616 | 83.50% | --- |
| Age (years) - Mean (SD) | 54.4 (17.0) |  | 58.2 (19.7) |  | 53.6 (16.4) |  | <0.001 |
| Age group (years) |  |  |  |  |  |  |  |
| <18 | 69 | 1.20% | 16 | 1.80% | 53 | 1.10% | <0.001 |
| 18-44 | 1486 | 26.90% | 208 | 22.80% | 1278 | 27.70% |  |
| 45-59 | 1619 | 29.30% | 182 | 19.90% | 1437 | 31.10% |  |
| 60-74 | 1731 | 31.30% | 301 | 33.00% | 1430 | 31.00% |  |
| ≥75 | 624 | 11.30% | 206 | 22.60% | 418 | 9.10% |  |
| **Sex** |  |  |  |  |  |  |  |
| Female | 1959 | 35.40% | 376 | 41.20% | 1583 | 34.30% | <0.001 |
| Male | 3570 | 64.60% | 537 | 58.80% | 3033 | 65.70% |  |
| **Length of stay (days) - Median (IQR)** | 7.0 (5.0-10.0) |  | 5.0 (2.0-8.0) |  | 7.4 (2.7-10.1) |  | <0.001 |
| **Length of stay categories (days)** |  |  |  |  |  |  |  |
| <5 | 1285 | 23.20% | 417 | 45.70% | 868 | 18.80% | <0.001 |
| 5-9 | 2865 | 51.80% | 317 | 34.70% | 2548 | 55.20% |  |
| 9-14 | 927 | 16.80% | 86 | 9.40% | 841 | 18.20% |  |
| ≥15 | 452 | 8.20% | 93 | 10.20% | 359 | 7.80% |  |
| **Ward type** |  |  |  |  |  |  |  |
| ICU | 1505 | 27.20% | 224 | 24.50% | 1281 | 27.80% | 0.046 |
| Ward | 4024 | 72.80% | 689 | 75.50% | 3335 | 72.20% |  |
| **Severity** |  |  |  |  |  |  |  |
| Mild | 3025 | 54.70% | 562 | 61.60% | 2463 | 53.40% | <0.001 |
| Moderate | 933 | 16.90% | 87 | 9.50% | 846 | 18.30% |  |
| Severe | 1571 | 28.40% | 264 | 28.90% | 1307 | 28.30% |  |
| **Diabetes** |  |  |  |  |  |  |  |
|  | 998 | 18.10% | 118 | 12.90% | 880 | 19.10% | <0.001 |
| **Hypertension** |  |  |  |  |  |  |  |
|  | 1364 | 24.70% | 170 | 18.60% | 1194 | 25.90% | <0.001 |
| **Dyslipidaemia** |  |  |  |  |  |  |  |
|  | 102 | 1.80% | 72 | 7.90% | 30 | 0.60% | <0.001 |
| **Hypothyroidism** |  |  |  |  |  |  |  |
|  | 405 | 7.30% | 137 | 15.00% | 268 | 5.80% | <0.001 |
| **Any other** |  |  |  |  |  |  |  |
|  | 193 | 3.50% | 11 | 1.20% | 182 | 3.90% | <0.001 |

### Number and duration of symptoms

Out of a total of 5529 cases in the two groups, only (78+464=) 542 (9.8%) reported no long-COVID symptom (Table 3). These were 8.5% of vaccinated and 10.1% of unvaccinated cases and the difference was not significant (P = 0.161). The other nearly 90% reported at least one long-COVID symptom. Mild cases at the time of admission reporting no long-COVID symptom were significantly (P = 0.010) less in the vaccinated (4.3%) compared with the unvaccinated (5.6%) group. The patients with moderate severity at admission with two or more long-COVID symptoms were less (1.9%) in vaccinated group compared with the unvaccinated group (5.8%) although the difference was not significant at 1% level (P = 0.018). There was no significant difference in the number of long-COVID symptoms between vaccinated and unvaccinated cases in those who recovered from severe form of COVID-19 (Table 3).

**Table 3.** Number of symptoms in cases with different severity of disease in the vaccinated and the unvaccinated cases.

| Severity | No. of symptoms | Vaccinated |  | Unvaccinated |  | P-value |
| --- | --- | --- | --- | --- | --- | --- |
|  |  | No. | Col % | No. | Col % |  |
| **Mild** | None | 39 | 4.3% | 259 | 5.6% | 0.010 |
|  | One | 336 | 36.8% | 1432 | 31.0% | 0.475 |
|  | Two+ | 187 | 20.5% | 772 | 16.7% | 0.375 |
|  | Total | 562 | 61.6% | 2463 | 53.4% |  |
| **Moderate** | None | 10 | 1.1% | 73 | 1.6% | 0.371 |
|  | One | 60 | 6.6% | 504 | 10.9% | 0.088 |
|  | Two+ | 17 | 1.9% | 269 | 5.8% | 0.018 |
|  | Total | 87 | 9.5% | 846 | 18.3% |  |
| **Severe** | None | 29 | 3.2% | 132 | 2.9% | 0.665 |
|  | One | 161 | 17.6% | 802 | 17.4% | 0.909 |
|  | Two+ | 74 | 8.1% | 373 | 8.1% | 0.867 |
|  | Total | 264 | 28.9% | 1307 | 28.3% |  |
| **Total** | None | 78 | 8.5% | 464 | 10.1% | 0.161 |
|  | One | 557 | 61.0% | 2738 | 59.3% | 0.341 |
|  | Two+ | 278 | 30.4% | 1414 | 30.6% | 0.912 |
|  | Total | 913 | 100.0% | 4616 | 100.0% |  |

As mentioned earlier, 542 (9.8%) patients reported no long-COVID symptoms at all. The remaining 90.2% of patients reported at least one symptom post-discharge. As many as 4104 out of a total of 5529 patients (74.2%) had symptoms that lasted for 1-4 weeks after discharge from hospital (short term PASC) (Table 4). During wave-3, out of 898 vaccinated patients, 671 (74.7%) had symptoms lasting between 1-4 weeks compared to 39 patients out of 65 unvaccinated cases (60%) (P=0.009). Unvaccinated in this wave generally had higher duration of symptoms. Similarly, if we compare symptom-weeks, it was observed that in wave-3 with omicron cases, significantly more vaccinated than unvaccinated patients reported 1-4 symptom-weeks (67.8% vs 49.2%, P=0.002) and non-significantly less patients with 5-11 symptom-week (10.6% vs 18.5%, P=0.051). Even in wave-2, 14 out of 15 patients (93.3%) in the vaccinated group versus 550 out of 767 in unvaccinated group (71.7%) had symptoms for 1-4 weeks after discharge from hospital, but this difference was not statistically significant (P=0.064).

**Table 4.**
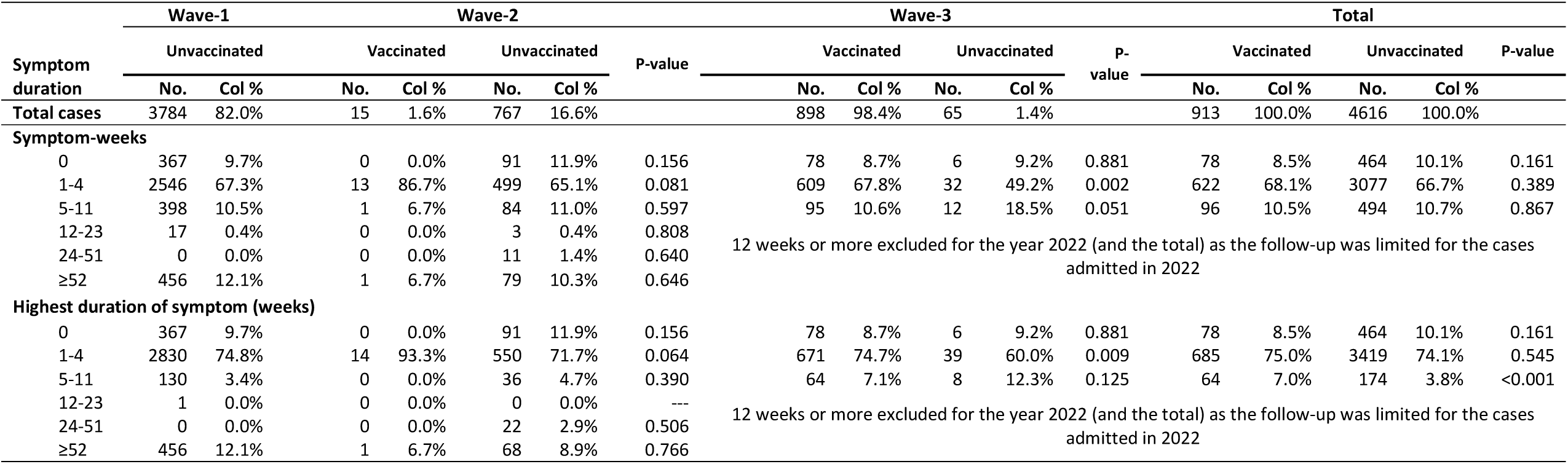
Symptom-weeks and highest duration of symptoms in the vaccinated and the unvaccinated groups.

Only 238 (4.3%) patients out of a total of 5529 continued to have symptoms for 1-3 months post only discharge and this number was not significantly different in the vaccinated and unvaccinated group. Only one patient of wave-1 had symptoms persisting between 3-6 months. The follow-up of those patients who continued to have symptoms for 6 months or more was restricted to only those patients who were admitted to the hospital during wave-1 and wave-2 (total numbers 4566). Of these, 547 (12.0%) reported symptoms more than 6 months duration. In this subgroup also, there was no significant difference between vaccinated and unvaccinated group. Significant number of patients continued to have long-COVID symptoms even beyond 52 weeks. This number was 456 patients out of a total of 2546 (12.1%) during wave-1, where all these patients were unvaccinated. During wave-2 (Delta), only one out of 15 vaccinated cases (6.7%) and 68 out of 767 unvaccinated cases (8.9%) continued to have long-COVID symptoms for more than 52 weeks; this difference was however, not statistically significant (P=0.76) (Table 4).

When the effect of wave, which also correspond to corona variant, age, sex, length of stay, severity and comorbidities is eliminated through multivariable logistic regression, the adjusted odds ratio (aOR = 1.27) for reporting of symptoms was higher in the vaccinated group but this failed to be statistically significant (P = 0.546) despite large sample (Table 5). Because of a large sample, we can safely conclude that the there was no overall difference between the two groups in the reporting of symptoms despite significant difference in their characteristics (Table 1) although the results of logistic regression show that hypertension and diabetes were more common in those who reported symptoms (P < 0.001) (Table 5). There were, however, differences in specific symptoms, as reported later in this communication.

**Table 5.**
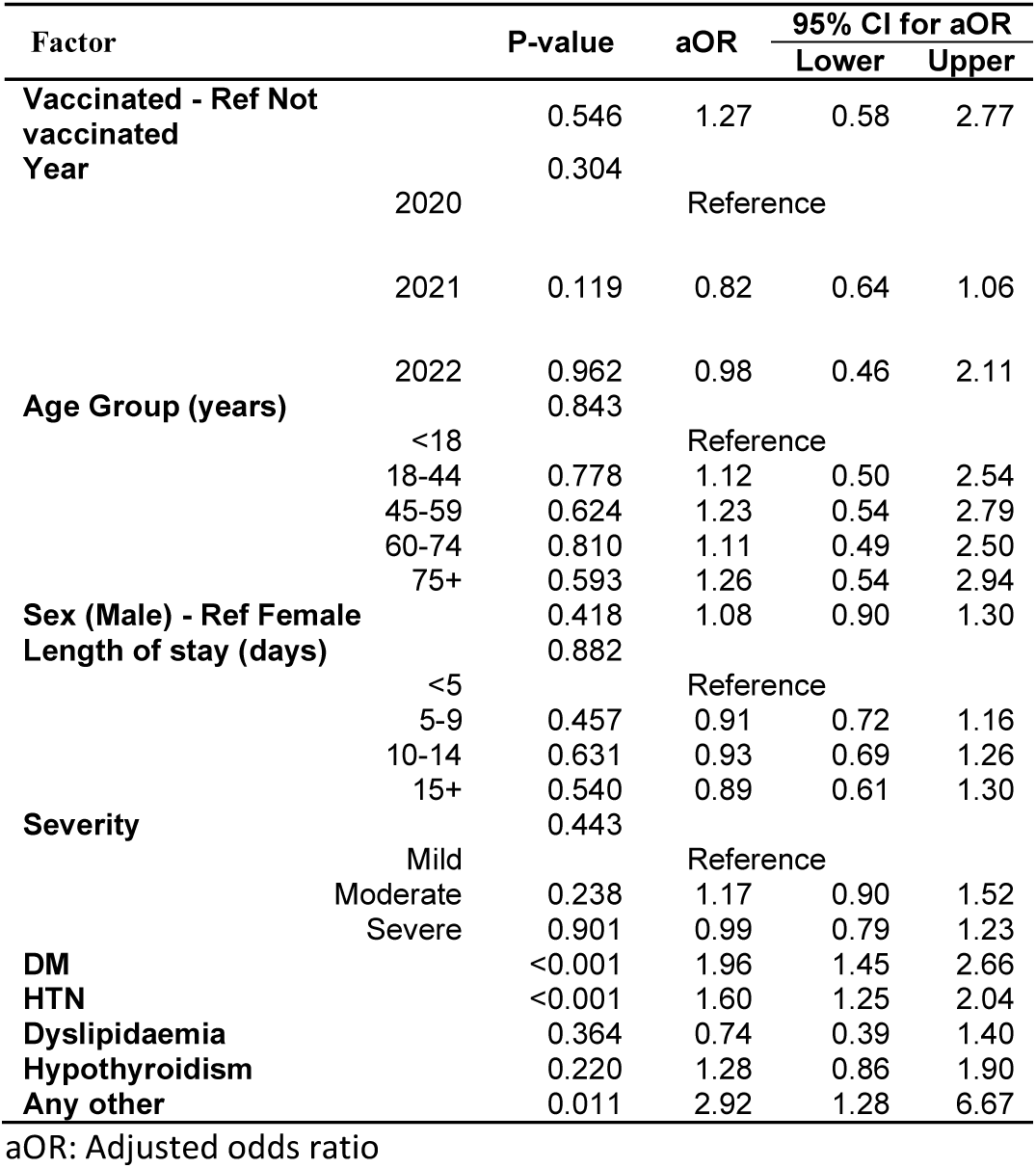
Results of the logistic regression of reporting of symptoms.

In some cases, symptoms were ongoing at the time of the survey. Thus, their duration is censored. Table 6 shows how symptom-weeks were affected by the covariates besides the vaccination status. No covariate had significant effect. The vaccination effect was also not significant (P = 0.424) on the symptom-weeks when the effect of the covariates is adjusted. Figure 2 shows that the ongoing symptoms drastically reduced after 5 symptom-weeks.

**Table 6.** Cox regression of symptom-weeks on vaccination groups and the covariates (age, sex, length of stay, severity, and comorbidities)

| Factor | P-value | HR | 95% CI for HR |  |
| --- | --- | --- | --- | --- |
|  |  |  | Lower | Upper |
| **Vaccinated – Ref Not vaccinated** | 0.424 | 0.97 | 0.89 | 1.05 |
| **Age Group (years)** | 0.303 |  |  |  |
| <18 |  | Reference |  |  |
| 18-44 | 0.263 | 1.17 | 0.89 | 1.53 |
| 45-59 | 0.256 | 1.17 | 0.89 | 1.53 |
| 60-74 | 0.471 | 1.10 | 0.84 | 1.44 |
| 75+ | 0.213 | 1.19 | 0.90 | 1.57 |
| **Sex (Male) – Ref Female** | 0.444 | 0.98 | 0.92 | 1.04 |
| **Length of stay (days)** | 0.282 |  |  |  |
| <5 |  | Reference |  |  |
| 5-9 | 0.209 | 0.95 | 0.89 | 1.03 |
| 10-14 | 0.728 | 1.02 | 0.93 | 1.12 |
| 15+ | 0.718 | 1.02 | 0.91 | 1.15 |
| **Severity** | 0.650 |  |  |  |
| Mild |  | Reference |  |  |
| Moderate | 0.359 | 0.96 | 0.89 | 1.04 |
| Severe | 0.672 | 0.99 | 0.92 | 1.06 |
| **DM** | 0.707 | 0.99 | 0.91 | 1.06 |
| **HTN** | 0.761 | 1.01 | 0.95 | 1.08 |
| **Dyslipidaemia** | 0.258 | 1.13 | 0.91 | 1.41 |
| **Hypothyroidism** | 0.061 | 0.90 | 0.80 | 1.01 |
| **Any other** | 0.701 | 1.03 | 0.88 | 1.21 |

HR = Hazard ratio. In this case, the 'hazard' of completing the duration (as opposed to continuing at the time of the survey) or getting rid of the symptoms by the time of the survey in vaccinated vs. unvaccinated group.

**Figure 2.**
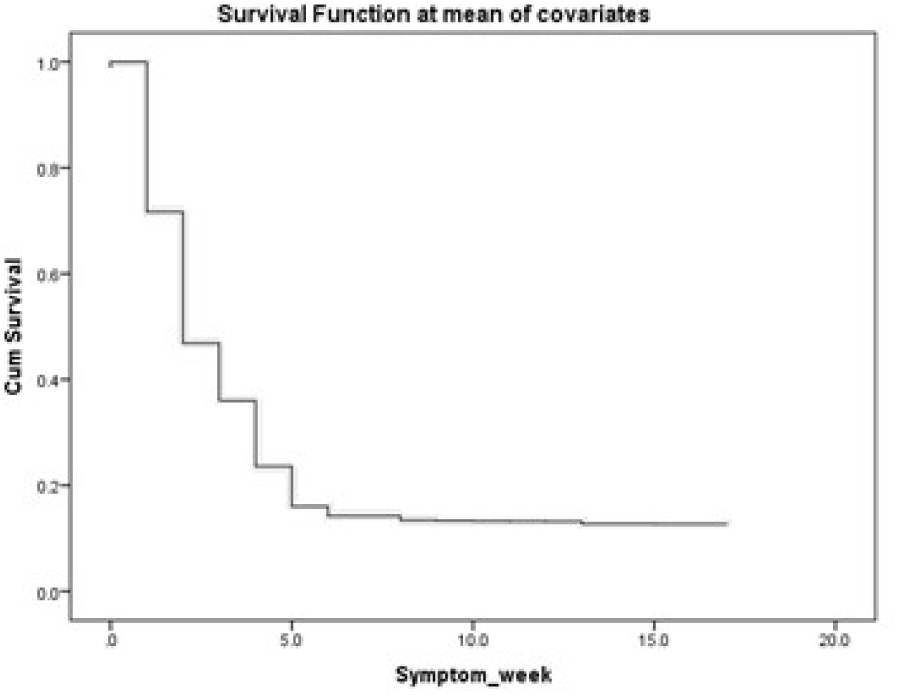
Probability of ongoing symptoms at different symptom-weeks (Cum Survival = probability of ongoing symptoms at the time of the survey)

### Specific symptoms

Fatigue, insomnia, and myalgia were the most commonly reported symptoms by both the groups. The symptoms that were significantly more often reported by vaccinated group were memory loss, cold, fever (five times), dyspnoea, eyesight (seven times), weight loss, headache (Table 7). The symptoms that were reported significantly less often in the vaccinated group were loss of taste, anxiety, memory loss, mental fogging, gastric issues, insomnia, myalgia, cough. Other symptoms were not affected by vaccination (Table 7).

**Table 7.** Comparison of vaccinated and unvaccinated for reporting of specific symptoms.

| Symptom | Group |  |  |  | P-value |
| --- | --- | --- | --- | --- | --- |
|  | Vaccinated |  | Not vaccinated |  |  |
|  | No. | Col% | No. | Col% |  |
| Total | 913 | 100.0% | 4616 | 100.0% |  |
| Loss of taste | 10 | 1.1% | 98 | 2.1% | 0.040 |
| Loss of smell | 4 | 0.4% | 25 | 0.5% | 0.692 |
| Low oxygen level | 2 | 0.2% | 18 | 0.4% | 0.432 |
| Anxiety | 28 | 3.1% | 260 | 5.6% | 0.001 |
| Memory loss | 17 | 1.9% | 37 | 0.8% | 0.003 |
| Low mood | 62 | 6.8% | 279 | 6.0% | 0.392 |
| Mental fogging/ lack of concentration | 16 | 1.8% | 172 | 3.7% | 0.003 |
| Vomiting | 63 | 6.9% | 259 | 5.6% | 0.128 |
| Gastric issues | 79 | 8.7% | 538 | 11.7% | 0.008 |
| Insomnia | 99 | 10.8% | 736 | 15.9% | <0.001 |
| Fatigue | 143 | 15.7% | 795 | 17.2% | 0.251 |
| Cold | 78 | 8.5% | 256 | 5.5% | 0.001 |
| Fever | 73 | 8.0% | 65 | 1.4% | <0.001 |
| Myalgia | 89 | 9.7% | 732 | 15.9% | <0.001 |
| Cough | 65 | 7.1% | 517 | 11.2% | <0.001 |
| Dyspnoea | 53 | 5.8% | 135 | 2.9% | <0.001 |
| Chest pain | 73 | 8.0% | 341 | 7.4% | 0.523 |
| Skin allergy | 43 | 4.7% | 215 | 4.7% | 0.946 |
| Eyesight | 35 | 3.8% | 24 | 0.5% | <0.001 |
| Weight loss | 50 | 5.5% | 87 | 1.9% | <0.001 |
| Hair loss | 14 | 1.5% | 59 | 1.3% | 0.537 |
| Headache/migraine | 90 | 9.9% | 194 | 4.2% | <0.001 |
| No symptom | 78 | 8.5% | 464 | 10.1% | 0.161 |

Significantly higher odds of reporting were observed for eyesight, fever, weight loss, headache, memory loss, dyspnoea, and cold in the vaccinated patients, and significantly lower odds of reporting of insomnia, cough, myalgia, anxiety, loss of taste (marginal), and mental fogging. The unadjusted odds ratios and their 95% CI are shown in the Figure 3 in deceasing order. After adjustment for the covariates (age, sex, length of stay, severity, and comorbidities), the aORs continued to be nearly the same as the unadjusted ORs, implying that the effect of the covariates on the reporting of symptoms in the vaccinated relative to the unvaccinated group was minimal (Table 8).

**Table 8.**
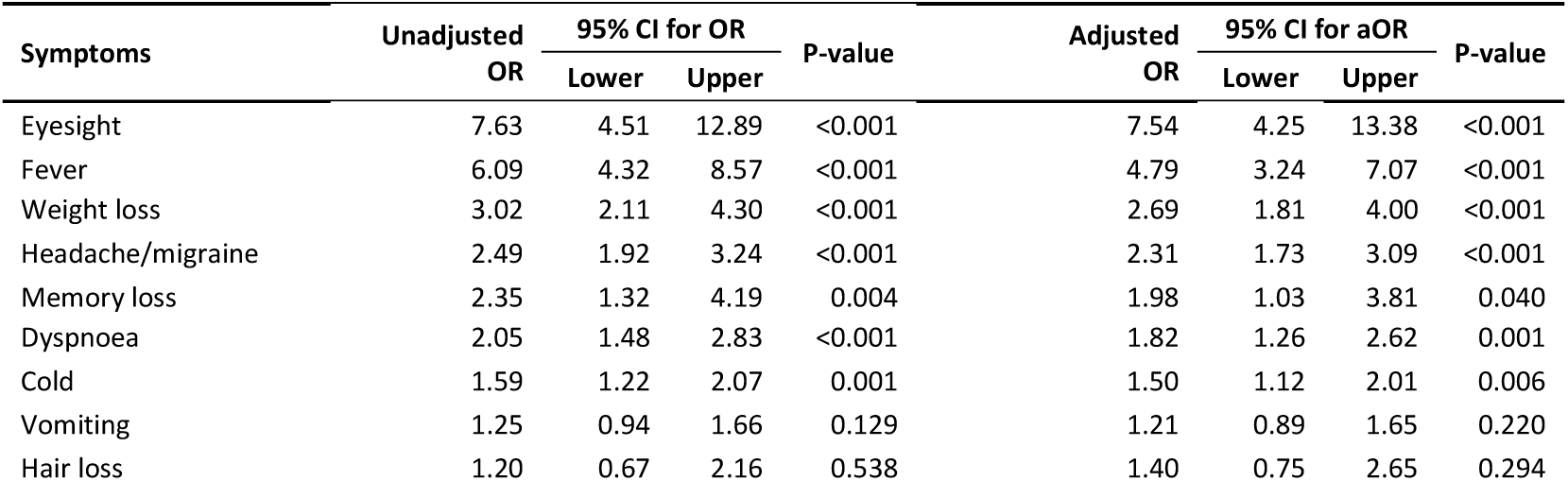
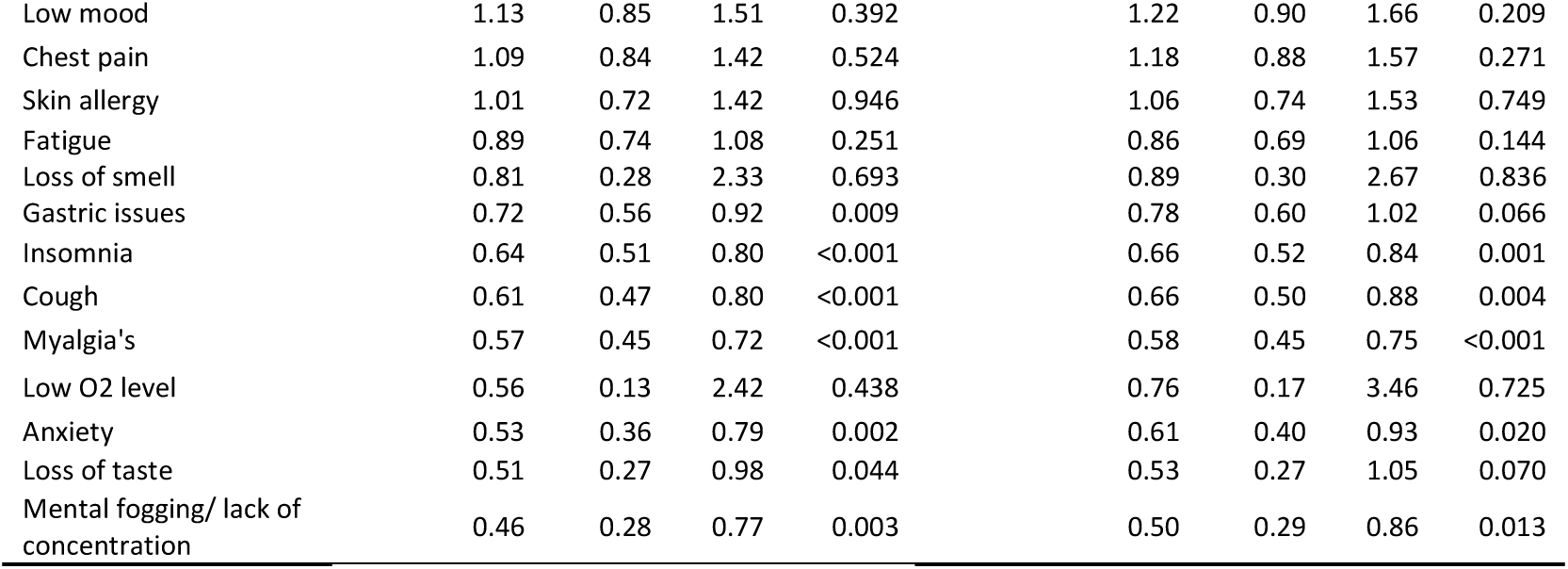
Odds ratio (in order from maximum to minimum) of reporting various symptoms in the vaccinated patients vs. the unvaccinated patients.

**Figure 3.**
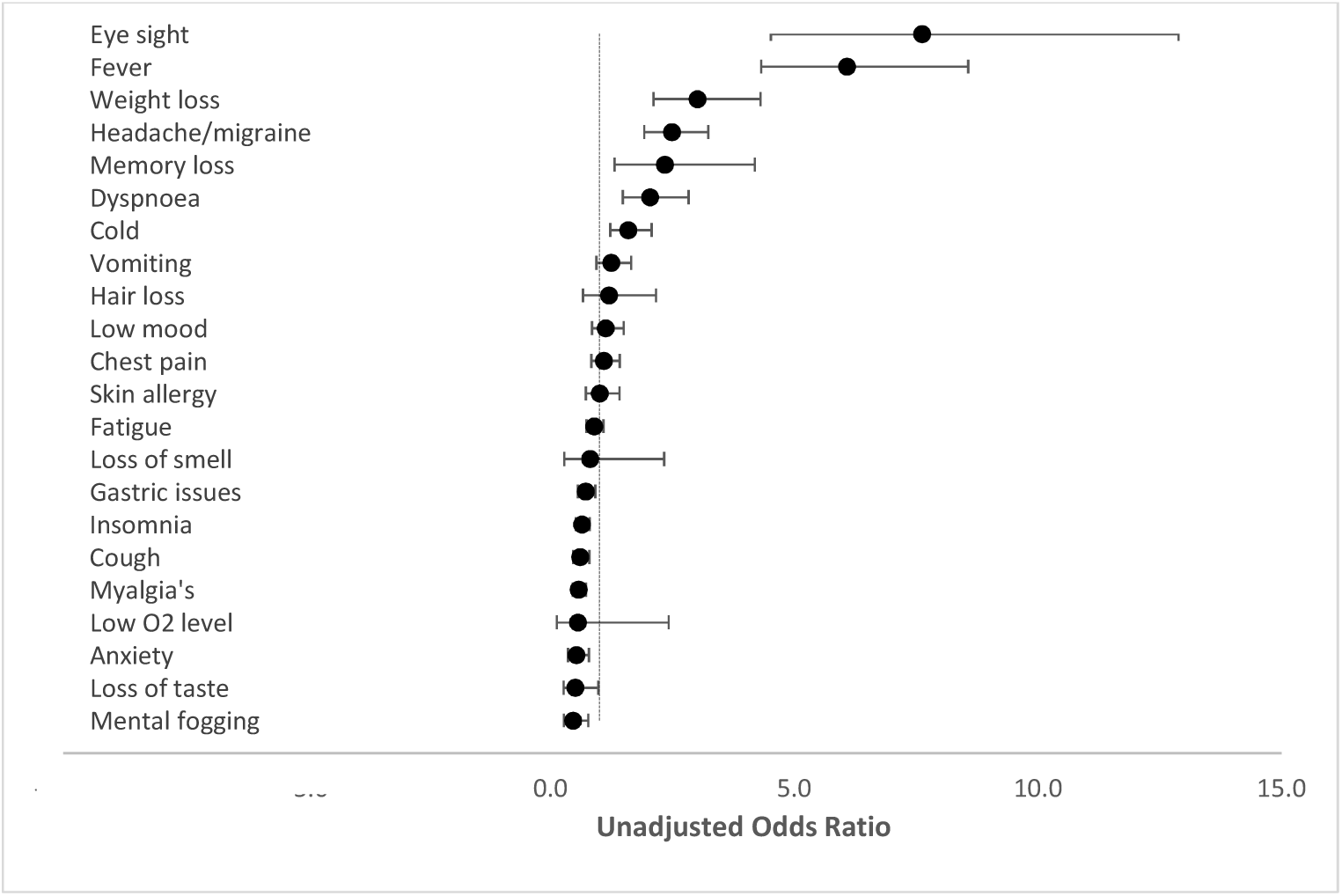
Odds ratio of reporting of specific symptoms by the vaccinated group vs. unvaccinated group.

The duration of symptoms is severely affected by the period of follow-up, and this period was widely different in vaccinated and unvaccinated cases. Thus, these two groups are not comparable for the duration of symptoms. P-values for difference between vaccinated and unvaccinated is not shown in the table as they are not valid in this case. However, the durations of different symptoms within each group are comparable. In the vaccinated cases, the longest duration symptoms on average were hair loss, memory loss, and loss of smell. In the unvaccinated cases also, the long duration symptoms were memory loss, weight loss, eyesight, and hair loss. Amongst other symptoms, gastric issue, loss of taste, and myalgia were the longest in both the groups (5 to 7 weeks in the vaccinated and 17 to 26 weeks in the unvaccinated because of longer follow-up). Had vaccinated group been followed up for longer duration, perhaps the durations would be the same. Despite shorter follow-up, the vaccinated group had significantly higher duration of low mood, mental fogging, vomiting, cold, fever, dyspnoea, and chest pain (Table 9).

**Table 9.** Mean and median duration of different symptoms in the cases in vaccinated and unvaccinated (*)

| Symptoms | Vaccinated |  |  | Unvaccinated |  |  |
| --- | --- | --- | --- | --- | --- | --- |
|  | No. | Duration (weeks) |  | No. | Duration (weeks) |  |
|  |  | Mean | Median |  | Mean | Median |
| **Cases with at least one symptom** | 835 | 3.8 | 2.0 | 4152 | 12.2 | 2.0 |
| Loss of taste | 10 | 7.2 | 5.0 | 98 | 20.0 | 5.0 |
| Loss of smell | 4 | 10.0 | 11.5 | 25 | 6.7 | 4.0 |
| Low O2 level | 2 | 3.0 | 3.0 | 18 | 3.1 | 3.0 |
| Anxiety | 28 | 2.8 | 2.0 | 260 | 2.7 | 2.0 |
| Memory loss | 17 | 10.2 | 11.0 | 37 | 44.8 | 55.0 |
| Low mood | 62 | 1.9 | 2.0 | 279 | 1.6 | 2.0 |
| Mental fogging/ lack of concentration | 16 | 4.4 | 2.0 | 172 | 1.5 | 1.0 |
| Vomiting | 63 | 1.3 | 1.0 | 259 | 1.0 | 1.0 |
| Gastric issues | 79 | 4.9 | 2.0 | 538 | 26.2 | 2.0 |
| Insomnia | 99 | 1.8 | 2.0 | 736 | 4.3 | 2.0 |
| Fatigue | 143 | 2.9 | 2.0 | 795 | 2.8 | 3.0 |
| Cold | 78 | 2.1 | 2.0 | 256 | 1.4 | 1.0 |
| Fever | 73 | 2.3 | 2.0 | 65 | 1.1 | 1.0 |
| Myalgia's | 89 | 5.2 | 3.0 | 732 | 17.1 | 3.0 |
| Cough | 65 | 2.0 | 2.0 | 517 | 2.9 | 2.0 |
| Dyspnoea | 53 | 3.9 | 3.0 | 135 | 1.6 | 1.0 |
| Chest pain | 73 | 1.9 | 2.0 | 341 | 1.3 | 1.0 |
| Skin allergy | 43 | 1.7 | 2.0 | 215 | 1.8 | 2.0 |
| Eyesight | 35 | 3.7 | 1.0 | 24 | 52.6 | 71.5 |
| Weight loss | 50 | 6.3 | 2.5 | 87 | 54.5 | 71.0 |
| Hair loss | 14 | 11.5 | 13.0 | 59 | 53.8 | 71.0 |
| Headache/migraine | 90 | 5.8 | 2.0 | 194 | 28.4 | 2.0 |

**\*542 cases with no symptom excluded**

### Wave comparison

Amongst the COVID-19 hospitalised patients during the three waves, there was a significantly (P< 0.001) higher percentage of younger (<18 years) patients in wave-3 (omicron) (3.9%) compared to those during wave-1 (0.6%) and wave-2 (0.9%) (Table 10). Similarly, there was also a significantly higher percentage of elderly (> 75 years) patients admitted in wave-3 (22.1%) compared to those admitted during wave-1 (9.0%) and wave-2 (8.5%) (P < 0.001). Significantly higher proportion of female patients (41.2%) were admitted in wave-3 compared to wave-1 (33.7%) and wave-2 (37.5%) (P<0.001). The mean length of stay in hospital was significantly shorter during wave-3 (6.6 days) compared to wave-1 (8.6 days) and wave-2 (7.8 days) (P<0.001). Nearly half (46.9%) of the admitted patients stayed for less than 5 days during wave-3 while this percentage during wave-1 was only 17.7% and during wave-2 22.3% (P<0.001). However, the percentage of patients going to ICU remained the same at around 25% across the waves. Overall, 61.6% of patients were clinically categorised as having mild disease during wave-3 hospitalisation, whereas these percentages during wave-1 were 54.9% and during wave-2 were 46.8% (P<0.001). Among comorbidities, it was observed that there were significantly lesser percentage of diabetics and hypertensive but more of dyslipidaemia and hypothyroidism during wave-3 compared to the earlier waves (P<0.001) (Table 10).

**Table 10:**
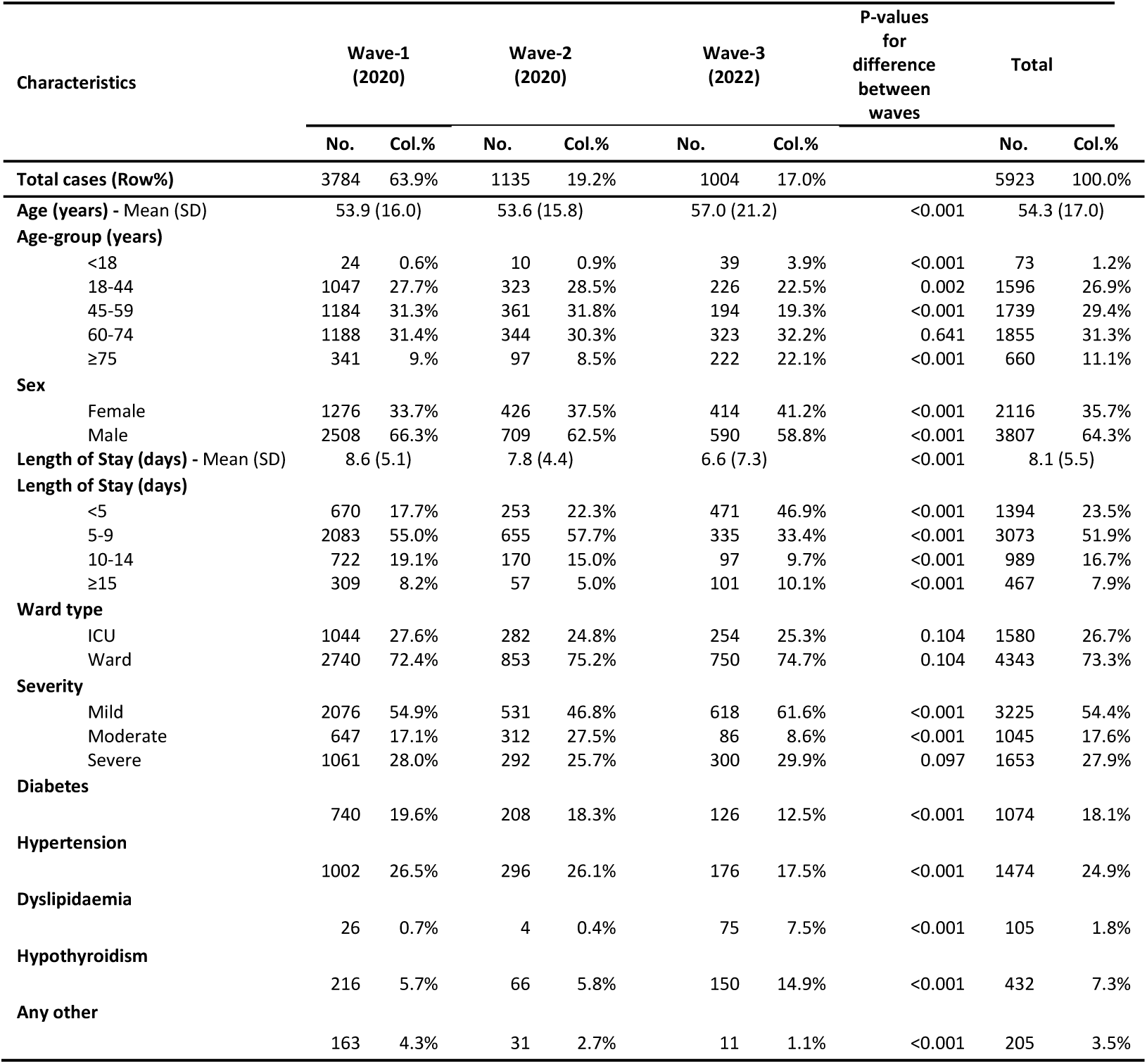
Year wise comparison of patient profiles.

| Characteristics | Wave-1<br>(2020) |  | Wave-2<br>(2020) |  | Wave-3<br>(2022) |  | P-values<br>for<br>difference<br>between<br>waves | Total |  |
| --- | --- | --- | --- | --- | --- | --- | --- | --- | --- |
|  | No. | Col.% | No. | Col.% | No. | Col.% |  | No. | Col.% |
| **Total cases (Row%)** | 3784 | 63.9% | 1135 | 19.2% | 1004 | 17.0% |  | 5923 | 100.0% |
| **Age (years) - Mean (SD)** | 53.9 (16.0) |  | 53.6 (15.8) |  | 57.0 (21.2) |  | <0.001 | 54.3 (17.0) |  |
| **Age-group (years)** |  |  |  |  |  |  |  |  |  |
| <18 | 24 | 0.6% | 10 | 0.9% | 39 | 3.9% | <0.001 | 73 | 1.2% |
| 18-44 | 1047 | 27.7% | 323 | 28.5% | 226 | 22.5% | 0.002 | 1596 | 26.9% |
| 45-59 | 1184 | 31.3% | 361 | 31.8% | 194 | 19.3% | <0.001 | 1739 | 29.4% |
| 60-74 | 1188 | 31.4% | 344 | 30.3% | 323 | 32.2% | 0.641 | 1855 | 31.3% |
| ≥75 | 341 | 9.0% | 97 | 8.5% | 222 | 22.1% | <0.001 | 660 | 11.1% |
| **Sex** |  |  |  |  |  |  |  |  |  |
| Female | 1276 | 33.7% | 426 | 37.5% | 414 | 41.2% | <0.001 | 2116 | 35.7% |
| Male | 2508 | 66.3% | 709 | 62.5% | 590 | 58.8% | <0.001 | 3807 | 64.3% |
| **Length of Stay (days) - Mean (SD)** | 8.6 (5.1) |  | 7.8 (4.4) |  | 6.6 (7.3) |  | <0.001 | 8.1 (5.5) |  |
| **Length of Stay (days)** |  |  |  |  |  |  |  |  |  |
| <5 | 670 | 17.7% | 253 | 22.3% | 471 | 46.9% | <0.001 | 1394 | 23.5% |
| 5-9 | 2083 | 55.0% | 655 | 57.7% | 335 | 33.4% | <0.001 | 3073 | 51.9% |
| 10-14 | 722 | 19.1% | 170 | 15.0% | 97 | 9.7% | <0.001 | 989 | 16.7% |
| ≥15 | 309 | 8.2% | 57 | 5.0% | 101 | 10.1% | <0.001 | 467 | 7.9% |
| **Ward type** |  |  |  |  |  |  |  |  |  |
| ICU | 1044 | 27.6% | 282 | 24.8% | 254 | 25.3% | 0.104 | 1580 | 26.7% |
| Ward | 2740 | 72.4% | 853 | 75.2% | 750 | 74.7% | 0.104 | 4343 | 73.3% |
| **Severity** |  |  |  |  |  |  |  |  |  |
| Mild | 2076 | 54.9% | 531 | 46.8% | 618 | 61.6% | <0.001 | 3225 | 54.4% |
| Moderate | 647 | 17.1% | 312 | 27.5% | 86 | 8.6% | <0.001 | 1045 | 17.6% |
| Severe | 1061 | 28.0% | 292 | 25.7% | 300 | 29.9% | 0.097 | 1653 | 27.9% |
| **Diabetes** | 740 | 19.6% | 208 | 18.3% | 126 | 12.5% | <0.001 | 1074 | 18.1% |
| **Hypertension** | 1002 | 26.5% | 296 | 26.1% | 176 | 17.5% | <0.001 | 1474 | 24.9% |
| **Dyslipidaemia** | 26 | 0.7% | 4 | 0.4% | 75 | 7.5% | <0.001 | 105 | 1.8% |
| **Hypothyroidism** | 216 | 5.7% | 66 | 5.8% | 150 | 14.9% | <0.001 | 432 | 7.3% |
| **Any other** | 163 | 4.3% | 31 | 2.7% | 11 | 1.1% | <0.001 | 205 | 3.5% |

When all the years are combined, the most commonly reported symptoms were fatigue (17.0%), followed by insomnia (15.1%) and myalgia (15.0%). The cases admitted in the third wave had relatively less of loss of taste, anxiety, mental fogging, insomnia, myalgia, and cough (Table 11).

**Table 11:** Comparison of symptoms in the patients admitted in different waves.

| Characteristics | Wave 1 |  | Wave 2 |  | Wave 3 |  | P-value for<br>the<br>difference<br>between the<br>years | Total |  |
| --- | --- | --- | --- | --- | --- | --- | --- | --- | --- |
|  | No. | Col.% | No. | Col.% | No. | Col.% |  | No. | Col.% |
| **Total cases (Row%)** | 3784 | 63.9% | 1135 | 19.2% | 1004 | 17.0% |  | 5923 | 100.0% |
| **No. of symptom** |  |  |  |  |  |  |  |  |  |
| None | 367 | 9.7% | 101 | 8.9% | 90 | 9.0% | 0.622 | 558 | 9.4% |
| One | 2243 | 59.3% | 703 | 61.9% | 603 | 60.1% | 0.274 | 3549 | 59.9% |
| Two+ | 1174 | 31.0% | 331 | 29.2% | 311 | 31.0% | 0.477 | 1816 | 30.7% |
| **Highest duration of the**<br>**symptom (week)** |  |  |  |  |  |  |  |  |  |
| 0 | 367 | 9.7% | 101 | 8.9% | 90 | 9.0% | 0.622 | 558 | 9.4% |
| 1-4 | 2830 | 74.8% | 844 | 74.4% | 739 | 73.6% | 0.741 | 4413 | 74.5% |
| 5-25 | 131 | 3.5% | 53 | 4.7% | 175 | 17.4% | <0.001 | 359 | 6.1% |
| 26-52 | 0 | 0.0% | 46 | 4.1% | 0 | 0.0% | <0.001 | 46 | 0.8% |
| ≥52 | 456 | 12.1% | 91 | 8.0% | 0 | 0.0% | <0.001 | 547 | 9.2% |
| **Specific symptom** |  |  |  |  |  |  |  |  |  |
| Loss of taste | 74 | 2.0% | 36 | 3.2% | 12 | 1.2% | 0.004 | 122 | 2.1% |
| Loss of Smell | 19 | 0.5% | 9 | 0.8% | 5 | 0.5% | 0.494 | 33 | 0.6% |
| Low O2 level | 16 | 0.4% | 3 | 0.3% | 2 | 0.2% | 0.485 | 21 | 0.4% |
| Anxiety | 209 | 5.5% | 76 | 6.7% | 31 | 3.1% | 0.001 | 316 | 5.3% |
| Memory loss | 26 | 0.7% | 10 | 0.9% | 20 | 2.0% | 0.001 | 56 | 0.9% |
| Low mood | 217 | 5.7% | 77 | 6.8% | 66 | 6.6% | 0.332 | 360 | 6.1% |
| Mental fogging/ lack of concentration | 146 | 3.9% | 35 | 3.1% | 17 | 1.7% | 0.003 | 198 | 3.3% |
| Vomiting | 220 | 5.8% | 71 | 6.3% | 67 | 6.7% | 0.565 | 358 | 6.0% |
| Gastric issues | 439 | 11.6% | 126 | 11.1% | 90 | 9.0% | 0.060 | 655 | 11.1% |
| Insomnia | 615 | 16.3% | 173 | 15.2% | 107 | 10.7% | <0.001 | 895 | 15.1% |
| Fatigue | 646 | 17.1% | 214 | 18.9% | 149 | 14.8% | 0.048 | 1009 | 17.0% |
| Cold | 209 | 5.5% | 51 | 4.5% | 90 | 9.0% | <0.001 | 350 | 5.9% |
| Fever | 51 | 1.3% | 16 | 1.4% | 80 | 8.0% | <0.001 | 147 | 2.5% |
| Myalgia's | 606 | 16.0% | 183 | 16.1% | 101 | 10.1% | <0.001 | 890 | 15.0% |
| Cough | 432 | 11.4% | 129 | 11.4% | 71 | 7.1% | <0.001 | 632 | 10.7% |
| Dyspnoea | 117 | 3.1% | 22 | 1.9% | 59 | 5.9% | <0.001 | 198 | 3.3% |
| Chest pain | 296 | 7.8% | 79 | 7.0% | 82 | 8.2% | 0.533 | 457 | 7.7% |
| Skin allergy | 182 | 4.8% | 43 | 3.8% | 49 | 4.9% | 0.326 | 274 | 4.6% |
| Eyesight | 21 | 0.6% | 1 | 0.1% | 39 | 3.9% | <0.001 | 61 | 1.0% |
| Weight loss | 68 | 1.8% | 18 | 1.6% | 56 | 5.6% | <0.001 | 142 | 2.4% |
| Hair loss | 49 | 1.3% | 15 | 1.3% | 15 | 1.5% | 0.887 | 79 | 1.3% |
| Headache/migraine | 162 | 4.3% | 42 | 3.7% | 99 | 9.9% | <0.001 | 303 | 5.1% |
| Any symptom | 3417 | 90.3% | 1034 | 91.1% | 914 | 91.0% | 0.622 | 5365 | 90.6% |
| No symptom | 367 | 9.7% | 101 | 8.9% | 90 | 9.0% | 0.622 | 558 | 9.4% |

### Vaccine comparison

We also analysed the variation in symptoms in the patients who received covaxin vs. those who received covishield. These were the two vaccines predominantly being used in India at that time. Only 26 (0.44%) patients reported some other or unknown vaccine. In our study, the patients in wave-1 were all unvaccinated, those from wave-2 were mixed population of completely vaccinated (15 / 782; 1.9%) and partially vaccinated / unvaccinated (767 / 782; 98.1%) while those from wave-3 were largely vaccinated group (898 / 963; 93.2%) and some partially vaccinated / unvaccinated (65 / 963; 6.7%).

Of those who were vaccinated, 73.6% received covishield. Detailed tables are omitted but we did not find any significant difference in long-COVID in the cases receiving covaxin and covishield. Those who took covishield had lower hospital stay with an average 5 days versus 6 days for covaxin; however, this was not statistically significant (P=0.071). But significantly (P = 0.017, marginal significance at 1% level) higher percentage of patients (47.9%) who received covishield stayed in hospital for less than 5 days compared to patients who got covaxin (38.9%). The cases receiving covaxin reported nearly similar symptoms and their duration except for vomiting which was more (P = 0.024) in the covishield group and skin allergy (P = 0.033) which was more in the covaxin group. These differences are not significant at 1% level.

## Discussion

### Effect of vaccine on COVID severity

Effectiveness of vaccination in reducing the severity of COVID-19 disease is now well established by various studies, including the need for hospitalization and death from COVID-19 [14,15]. Our study was conducted in patients who were admitted in hospital for COVID-19 over the three waves and in these patients also, the vaccinated group had a significantly shorter average length of stay compared to the unvaccinated group (5.0 vs 7.4 days) (P < 0.001). Significantly more patients had mild COVID-19 at the time of admission in the vaccinated group (61.6%) compared to the unvaccinated group (53.4%) (P < 0.001). Being tertiary care hospitals, ICU occupancy was higher due to direct referrals of sicker patients and ICU admissions were not significantly different in the two groups.**Effect of vaccine on occurrence of long-COVID**

There is no universally accepted definition of long-COVID. The National Institute for Health & Care Excellence (NICE) [1] defined long-COVID as signs and symptoms that continue or develop after acute COVID-19, including both ongoing symptomatic COVID-19 (from 4 to12 weeks) and post-COVID-19 syndrome (>12 weeks). The Centres for Disease Control and Prevention (CDC) defined long-COVID as a post COVID condition with a wide range of new, returning or ongoing health problems people experience four or more weeks after first being infected with the virus that causes COVID-19 [16]. WHO, in Oct 2021, defined long-COVID as a condition that occurs in individuals with a history of probable or confirmed (SARS-COV-2) infection, usually three months from the onset of COVID-19, with symptoms that last for at least two months and cannot be explained by an alternative diagnosis [17]. Most of the studies included in the review by UK Health Security Agency used different definitions of long-COVID [18]. In the present study, we have taken symptoms persisting or developing after discharge from the hospital as long-COVID.

In our study, nearly 9% of the patients reported at least one symptom after discharge from hospital. Several recent studies, including systematic reviews, have shown that around 5% to 87% of hospitalized patients experienced at least one or more post-COVID-19 symptoms for several weeks after discharge from hospital [19-23]. An Indian study on 773 patients by Senjam et al. [7] reported overall incidence of 33% with short-term long-COVID symptoms (4 to 12 weeks) and 13% with long-term long-COVID symptoms (≥ 12 weeks). The study by Kuodi et al. [7] from Israel reported 35% of their patients experienced long-COVID. Our rates at 9% are higher. The reason for this could be that we have included and not restricted to the criteria of > 4 weeks from the time of COVID-19 diagnosis. Our duration is from the discharge and not diagnosis. If the patients with symptoms of duration 4 weeks or less are excluded, the incidence in our cases too steeply declines to 15%. In our series,8.5% of vaccinated and 10.1% of unvaccinated cases reported no symptoms; this difference was not statistically significant (P = 0.161). However, in the mild COVID-19 group, significantly higher number of unvaccinated patients had no long-COVID symptoms (5.6%) compared to the vaccinated group (4.3%) (P = 0.010).

UK Health Security Agency [18] published a rapid evidence briefing on the effectiveness of vaccination against long-COVID. In their analysis, six of the 8 studies suggested that vaccinated cases were less likely to develop symptoms of long-COVID in the short term (within 4 weeks after infection), medium term (12 to 20 weeks after infection) and long term (6 months after infection) [4-8]. Antonelli et al. [28] found that fully vaccinated patients were almost half as likely to have symptoms lasting ≥ 28 days than unvaccinated patient (OR = 0.51, 95% CI: 0.32 to 0.82, p = 0.005).

Al–Aly et al. [4] reported in their retrospective cohort study that vaccinated cases were less likely to have at least one post-acute sequelae of COVID-19 at 6 months compared with the unvaccinated cases (HR = 0.87, 95% CI: 0.83 to 0.92). A retrospective cohort study by Herman et al. [5] found that fully vaccinated patients were less likely to develop of olfactory dysfunction after infection than unvaccinated patients (OR = 0.31, 95% CI: 0.10 to 0.94), but there was little evidence for an association between full vaccination and olfactory dysfunction at 4 weeks after end of infection (P = 0.59). An Israeli study by Koudi et al. [6] found that those with 2 or 3 doses of vaccine were 54% to 83% less likely to report 7 of the 10 most commonly reported symptoms than the unvaccinated. Senjam et al. [7] from India reported that fully vaccinated patients were less likely to have long-COVID symptoms than unvaccinated patients (OR = 0.55, 95% CI: 0.37 to 0.85). A retrospective study by Simon et al. [8] revealed that patients who received at least one dose of any of the three COVID vaccines in the USA prior to their diagnosis with COVID-19 were 7-10 times less likely to report two or more long-COVID symptoms compared to the unvaccinated patients.

Another matched case-control study by Taquet et al. [9] from the USA included 9,479 vaccinated and a similar number of unvaccinated cases who were followed up for 6 months after infection to study a composite long-COVID outcome. The study found no association between vaccination and composite long-COVID outcome in 6 months after infection with hazard ratio (HR) = 1.00 (95% CI: 0.95 to 1.06). In their study, 64.9% and 65.5% of vaccinated and unvaccinated patients had long-COVID respectively.

In our study, the results of logistic regression show that hypertension and diabetes were more common in those who reported long-COVID symptoms (P < 0.001). The study by Senjam et al. [7] also indicated a strong association between the pre-existing comorbidity and the presence of post COVID sequelae. Pre-existing co-morbidity such as hypertension, chronic respiratory diseases, diabetes mellitus were shown to be associated with the prolonged COVID-19 symptoms in other studies also [24,25,26].

### Effect of vaccine on duration of long-COVID

Groff et al. [2] identified 57 studies with 250,351 survivors of COVID-19, of which 197,777 (79%) were hospitalized during acute COVID-19. More than half of these experienced PASC 6 months after recovery. They divided their patients into 3 categories of symptom duration: Short term PASC: up to 1 month (median - IQR 54% - 45% to 69%; 13 studies). Intermediate term PASC: 2-5 months (median - IQR 55% - 34.8% to 65.5%; 38 studies). Long-term PASC: ≥ 6 months (median - IQR 54% - 31% to 67%; 9 studies). These categorizations were based on literature reports proposing a framework of COVID-19 infection progresses from an acute infection lasting approximately 2 weeks into a post-acute hyper-inflammatory illness lasting approximately 4 weeks, until ultimately entering late sequelae [19,27].

%%In our study, 74.7% of the vaccinated patients versus 6% of the unvaccinated patients reported symptoms that lasted 1-4 weeks after discharge from the hospital (short term PASC) (P=0.009) during the third wave. Although a similar trend was observed during wave-2, the difference was not statistically significant. A retrospective cohort study by Arjun et al. [10] from India, involving 487 patients, showed that fully vaccinated participants were more likely to have long-COVID symptoms 4 weeks from the date of diagnosis compared to unvaccinated patients (OR = 2.32, 95% CI: 1.17 to 4.58, P = 0.01).

In our study, we did not find any significant difference in the occurrence of intermediate and long term PASC between vaccinated and unvaccinated group. Almost 11.4% of the patients during the first two waves continued to have long-COVID symptoms beyond 52 weeks and there was no significant difference between the vaccinated and the unvaccinated group.

### Effect of vaccine on occurrence of specific symptoms of long-COVID

When the effect of wave, age, gender, length of hospital stay, severity of COVID-19 and comorbidities is eliminated through multivariate logistic regression, the adjusted odds ratio (aOR = 0.78) for reporting of symptoms was statistically not significant between vaccinated and unvaccinated group in our cases. When all the three waves and both groups (vaccinated and unvaccinated) are combined, the most commonly reported long-COVID symptoms in our study were fatigue (17.0%), followed by insomnia (15.1%) and myalgia (15.0%). In a study by Senjam et al. [7], the most commonly reported long-COVID symptoms were fatigue, pain in the joints and muscle, hair loss, headache, cough, breathlessness, sleep disorders, sore throat and decrease in smell and taste. These are somewhat different from what we observed.

In the vaccinated group, we found significantly higher odds of reporting eyesight issues (aOR = 7.54, 95% CI 4.25 to 13.38, P < 0.01). In a study by Senjam et al. [7], some patients experienced both near and distance visual impairment and dry eyes. They surmised these eye symptoms could be side effects of medication such as hydroxychloroquine that was commonly used during their acute management. Since the post-COVID assessment was done four or more weeks after recovery, symptoms like red and painful eyes could be due to immunological mechanisms.

Kuodi et al. [6], compared participants with 2 or 3 doses of vaccine with the unvaccinated participants, and observed relative risks less than 1 indicating that the vaccinated had less of fatigue, headache, weakness in arms and legs, persistent muscle pain, loss of concentration, hair loss, sleeping problems, dizziness, persistent cough, shortness of breath and more feeling fully recovered from COVID-19. The study by Taquet et al. [9] found no association between vaccination and composite long-COVID. Patients with 2 doses of vaccine were found to be less likely to be diagnosed with anosmia, fatigue, hair loss, intestinal lung disease, myalgia, and any other pain. Our findings suggest that vaccinated group had significantly higher odds of reporting eyesight issues, fever, weight loss, headache, memory loss, dyspnoea, cold and a significantly lower odds of reporting insomnia, cough, myalgia, and anxiety.

### Comparison between different waves

In India, wave-1 during 2020 was reported to be predominantly caused by the Wuhan strain of SARS-COV-2, while in wave-2 in the first half of 2021, the Delta strain was most prevalent. In wave-3 (Dec 2021 till Feb 2021) the Omicron strain caused majority of cases of COVID-19.

We observed no significant effect of the wave on the overall incidence of long-COVID. This could be interpreted that long-COVID had no correlation with the strain. The symptoms reported with less frequency during the third wave were loss of taste, anxiety, mental fogging, insomnia, and cough but certain symptoms had higher frequency such as memory loss, cold, dyspnea, ey sight issues and headache. These cases were mostly vaccinated. Further studies are needed to answer whether this difference is due to effect of vaccination or the viral strain.

We had significantly higher percentage of younger (<18 years) and elderly (≥75 years) patients in wave-3 compared to the previous two waves (P < 0.001). Significantly more female patients were admitted in wave-3 (41.2%) compared to wave-2 (37.5%) and wave-1 (33.7%) (P < 0.001). The mean length of stay in hospital was significantly shorter in wave-3 (6.6 days) compared to that during wave-2 (7.8 days) and wave-1 (8.6 days) (P < 0.001).

The disease was overall milder in wave-3 (61.6% of patients were categorized as having mild disease) compared to wave-2 (46.8%) and wave-1 (54.9%) (P < 0.001). It was also observed that there were significantly fewer diabetes and hypertension but more of dyslipidemia and hypothyroid patients during wave-3 compared to earlier waves (P < 0.001).

Antonelli et al. [28] reported that Omicron appears to cause less severe acute illness than previous variants, at least in the vaccinated subjects. They conducted a case-control observational study to identify the relative odds of long-COVID (symptoms > 4 weeks from start of COVID-19) in the UK during Omicron period compared with Delta period. Their results showed that among Omicron cases, 2501 (4.5%) of 56,003 people experienced long-COVID and, among Delta cases, 4469 (10.8%) of 41,361 people experienced long-COVID. They concluded that Omicron cases were less likely to experience long-COVID with an odds ratio ranging from 0.24 (0.20 – 0.32) to 0.50 (0.43 – 0.59).

In our study, 133 patients died in the post COVID period (from discharge to the time of tele-calling). The cause of death and their vaccination status was not assessed for this group. Al-Aly et al. [4] noted that people who survived the first 30 days of breakthrough COVID-19 exhibited an increased risk of death (HR 1.53; 95% CI: 1.36 to 1.72) and the excess burden of death was estimated at 11.45 (95% CI: 7.77 to 15.58) per 1000 persons with breakthrough COVID-19 at 6 months compared to the control group.

### Effect of vaccine type

In our study, 239 (26.4%) out of 907 vaccinated patients received covaxin and 668 (73.6%) received covishield vaccine. There was no significant difference in the demographic profile between those who got covaxin versus covishield. The patients who received covaxin reported nearly similar long-COVID symptoms and their duration, except for vomiting significantly (P = 0.024) more in covishield group and skin allergy (P = 0.033) significantly more in covaxin group. A retrospective study by Arjun et al. [10] included 487 patients, 59% of whom had 2 doses of COVID-19 vaccine (the majority had Covaxin) and they actually reported that fully vaccinated patients were more likely to have long-COVID symptoms at 4 weeks from the diagnosis. In a study by Taquet et al [9], 65% of the patients who were vaccinated had the Pfizer vaccine, 9% had Moderna vaccine, 1.6% had the Janssen vaccine, and 24% had an unspecified vaccine. They did not compare the incidence of long-COVID in different vaccine groups.

## Conclusion

This may be the first large scale study in India with upto a 2-year follow-up of hospitalized COVID-19 cases for the effect of vaccination on long-COVID symptoms. This may also be the first study to use “symptom-weeks” that considers both the number of symptoms and duration of symptoms, that may have combined effect on a patient’s overall quality of life.

We found that the fully vaccinated cases had reduced stay in the hospital and moderate severity cases were lower and milder COVID-19 cases higher. Thus, it seems that the vaccination helped in modifying the disease from moderate to mild in some cases. The proportion of severe cases remained unaffected.

Nearly 90% of COVID-19 patients reported at least one long-COVID symptom irrespective of their vaccination status but almost three-fourth of these had symptoms lasting up to a month. Nearly 11% reported symptoms even after one year. Interestingly, during wave-3, significantly more vaccinated patients reported short-term post-acute sequelae of COVID-19 than did the unvaccinated group. Most of the patients either had short term or long term PASC symptoms and a very few reported intermediate duration PASC symptoms. The cases with diabetes and hypertension had higher odds of reporting at least one symptom when the effect of vaccination, age, sex, severity, and length of stay was adjusted.

Most common symptoms reported by both the groups were fatigue, insomnia, and myalgia. Some symptoms, such as eyesight issues, fever, weight loss, headache, memory loss, dyspnoea and cold were more common in the vaccinated and some other such as insomnia, cough, and myalgia in the unvaccinated. Overall vaccination does not seem to significantly alter either the incidence or duration of long-COVID.

## Limitations

The present study included only those COVID-19 patients who were admitted in hospitals. Hence, it does not represent the overall burden of the long-COVID in a community. The symptoms of long-COVID were self-reported although a structured questionnaire may have reduced the error. The follow-up of wave-3 patients was up to a maximum period of 12 weeks only. Hence, the percentage of cases who would have continued to have symptoms have been missed in the present analysis. The definition of long-COVID is still not fully standardised and hence comparison between various studies becomes difficult and at times erroneous.

## Role of the Authors

SB designed the study concept, contributed patients for the study and wrote the manuscript; AI did the statistical analysis and review of document and MM contributed patients and assisted in writing and reviewing the manuscript.

## Contributions

We wish to acknowledge and thank the contribution made by Taruna Sharma, Rajesh Saxena, Bhawna Sharma and Cheshtha Kaila.

